# Persistence of a SARS-CoV-2 variant with a frameshifting deletion for the duration of a major outbreak

**DOI:** 10.1101/2022.09.06.22279658

**Authors:** Charles S.P. Foster, Rowena A. Bull, Nicodemus Tedla, Fernando Santiago, David Agapiou, Anurag Adhikari, Gregory J. Walker, Lok Bahadur Shrestha, Sebastiaan J. van Hal, Ki Wook Kim, William D. Rawlinson

## Abstract

Australia experienced widespread COVID-19 outbreaks from infection with the SARS-CoV-2 Delta variant between June 2021 and February 2022. Whole-genome sequencing of virus from an early case revealed a sub-consensus level of sequencing reads supporting a 17-nucleotide frameshift-inducing deletion in ORF7a that truncated the peptide sequence. The variant rapidly became represented at the consensus level (Delta-ORF7a^Δ17del^) in most of the outbreak cases in Australia. Retrospective analysis of ORF7a deletions in all GISAID clade GK Delta genomes showed that of 4,018,216 genomes, 134,751 (∼3.35%) possessed a deletion in ORF7a, with the ORF7a^Δ17del^ mutation by far the most common. Approximately 99.05% of Delta-ORF7a^Δ17del^ genomes on GISAID originated from the Australian Delta outbreak, and comprised 87% of genomes in the outbreak. *In vitro* comparison of lineages in cell culture showed a significantly greater proportion of cells were infected with Delta-ORF7a^Δ17del^ than with a contemporaneous Delta variant without ORF7a^Δ17del^ (Delta-ORF7a^intact^), and the proportion was also measurably higher than an early SARS-CoV-2 strain (A.2.2). These results showed that Delta-ORF7a^Δ17del^ potentially has a slight growth advantage compared to Delta-ORF7a^intact^. Delta-ORF7a^Δ17del^ viruses still produced ORF7a protein, but significantly less than A.2.2, in a different cellular distribution with a more diffuse expression throughout the cytoplasm of infected cells. These data suggest that the proliferation of Delta-ORF7a^Δ17del^ genomes during the Australian Delta outbreak was likely not a result of an intrinsic benefit of the ORF7a^Δ17del^ mutation, but rather a chance founder effect. Nonetheless, the abundance of different ORF7a deletions in genomes worldwide suggests these have some benefit to virus transmission.

**IMPORTANCE:** Deletions in the ORF7a region of SARS-CoV-2 have been noted since early in the COVID-19 pandemic, but are generally reported as transient mutations that are quickly lost in the population. Consequently, ORF7a deletions are considered disadvantageous to the virus through possible loss-of-function effects. In constrast to these earlier reports, we present the first report of a SARS-CoV-2 variant with an ORF7a deletion that dominated for the entirety of a protracted outbreak, and found no associated fitness disadvantage or advantage in cell culture. The relatively common rise and fall of ORF7a deletion variants over time likely represent chance founder events followed by proliferation until a more fit variant(s) is introduced to the population. Our global clade-level survey of ORF7a deletions will be a useful resource for future studies into this gene region.

## Introduction

ORF7a is one of seven non-structural proteins encoded by SARS-CoV-2, which normally localise to the Golgi, endoplasmic reticulum (ER) and cell surface (1). Of these seven non-structural proteins, three including ORF7a (ORF3b, ORF6, ORF7a) are reported to modulate antiviral responses through processes including reduction of type 1 interferon (2). The ORF7a region of SARS-CoV-2 is an ortholog of the corresponding ORF7a region of SARS-CoV, which has previously been documented to be an antagonist of host restriction factor BST2/CD317/Tetherin (3, 4). Host BST2 has been reported to induce inhibit viral infection by promoting apoptosis of infected cells (5). During SARS-CoV-2 infections, this apoptotic pathway is inhibited by ORF7a cellular concentrations, resulting in increased SARS-CoV-2 replication and overall more virus being released (4). Additionally, the ORF7a accessory protein of SARS-CoV-2 has been linked to modulation of inflammatory responses via its ability to bind CD14+ monocytes and drive a proinflammatory response (6).

Despite the role of ORF7a in immune evasion, deletions at the C-terminus of ORF7a in SARS-CoV-2 have been detected since the early stages of the COVID-19 pandemic. One of the earliest ORF7a deletions, reported on in March 2020, was an 81-nucleotide in-frame deletion that removed the signal peptide and beta strand sequences of the ORF7a protein (3). In July 2020, Nemudryi et al. (2) detected a 115-nt disruptive deletion in ORF7a, and showed that that this mutation altered host immune responses by enhanced interferon signalling. Placing this sequence in a global context, 189 unique ORF7a variants were detected from the ∼181,000 genomes uploaded to GISAID at the time. These reported variants were also found to be transient, with few instances of transmission within a host population (2). This may be due to reduced viral fitness when ORF7a function is altered through in-frame or disruptive mutations, possibly as a consequence of reduced suppression of host innate immunity (2).

For the first 18 months of the COVID-19 pandemic, there were relatively few SARS-CoV-2 infections in the Australian community compared to global case numbers, particularly in New South Wales (NSW). The first widespread cases of COVID-19 in NSW began in June 2021, stemming from community transmission of the SARS-CoV-2 Delta variant (initially B.1.617.2, later designated as AY.^*^ Pango sublineages). Outbreak cases soon spread to other Australian states, leading to widespread community transmission. This ‘Delta outbreak’ spanned from mid-June 2021 until mid-February 2022 The first virus of the Delta outbreak was collected on June 16^th^ 2021, and the consensus genome exhibited a 100% match, including a full-length intact ORF7a gene (Delta-ORF7a^intact^), when compared to a USA consensus sequence, the likely progenitor of the Australian outbreak (Figure 1A). However, routine sequencing of subsequent cases identified a 17-nt deletion in the ORF7a gene spanning genome positions 27607–27623 (Figure 1). On further analysis of the first sequence, a fraction of sequencing reads (∼25%) supported this 17-nt deletion (Delta-ORF7a^Δ17del^), suggesting a mixed infection in the index case with onward transmission of both viral ‘subtypes’ ^(7)^. The Delta outbreak cases soon grew to be characterised by increased frequency of detection of Delta-ORF7a^Δ17del^.

**Figure 1.**
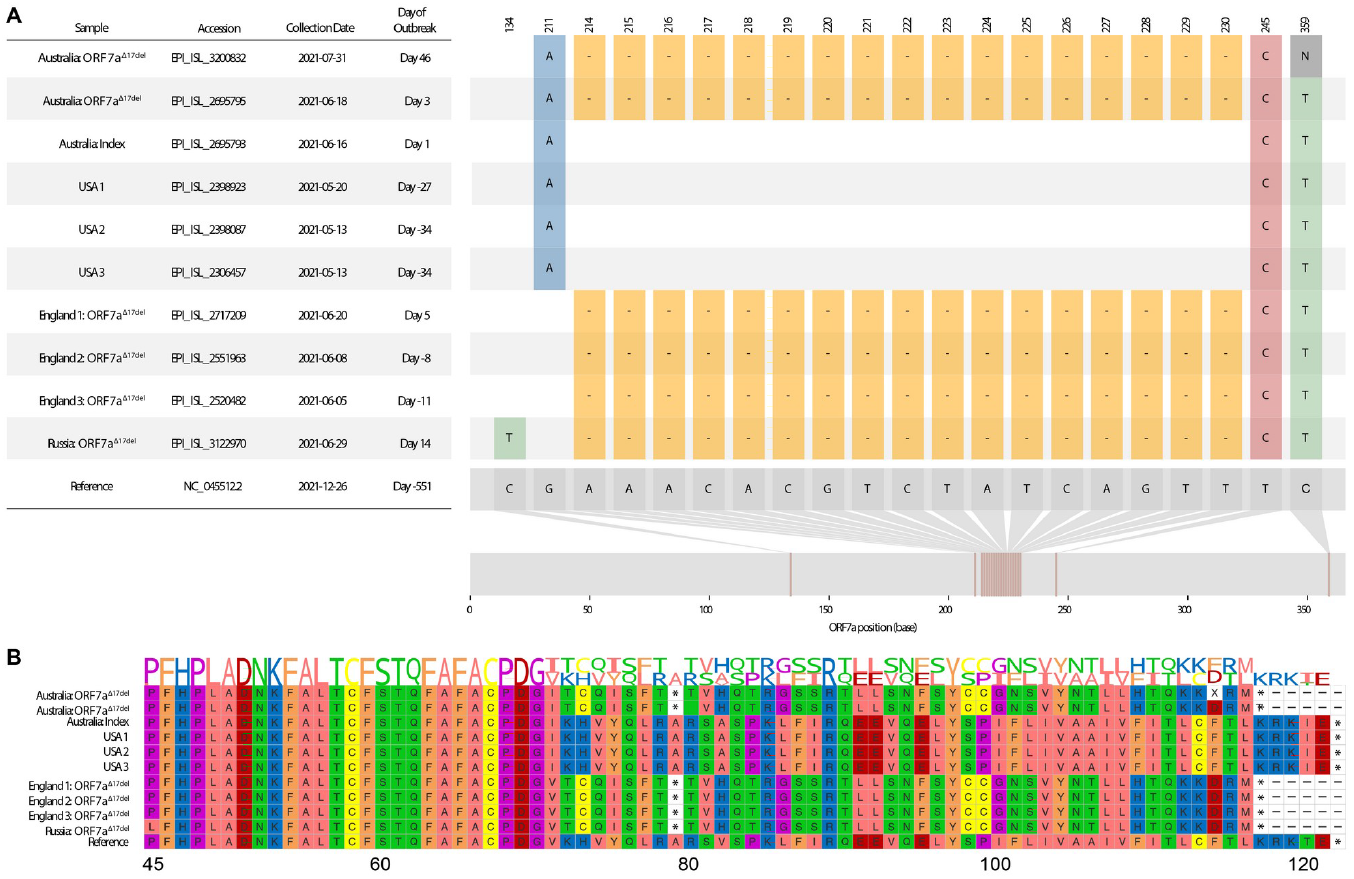
Visualisation of a 17-nucleotide deletion in ORF7a of Delta variant SARS-CoV-2 sequences (ORF7a^Δ17del^), and the corresponding consequences at the peptide level. Sequences chosen for visualisation include the index case of the Australian Delta outbreak (lacking ORF7a^Δ17del^), the first Australian case possessing ORF7a^Δ17del^ during the outbreak period, an Australian sample from ∼1.5 months into the outbreak that possesses ORF7a ^17del^, the three USA samples (all lacking ORF7a^Δ17del^) that match the Australian index case, and a selection of four non-Australian samples to possess ORF7a^Δ17del^, some of which were collected before the Australian outbreak had begun. **A**. Sample dates of all chosen samples relative to the index case of the Australian Delta outbreak (16 June 2021), and all variants in the sample genomes relative to the SARS-CoV-2 reference genome (created in part using snipit (https://github.com/aineniamh/snipit). **B**. The consequences of ORF7a^Δ17del^ on the amino acid translation of ORF7a, leading to a premature stop codon after amino acid 78 and a truncated peptide sequence (∼64% complete). Amino acids 1–44 are identical to the SARS-CoV-2 reference genome in all samples and are not presented. Figure adapted from (7).

We undertook this retrospective study to better understand the spread of Delta-ORF7a^Δ17del^, and explore possible reasons why, unlike other ORF7a variants (2), Delta-ORF7a^Δ17del^ persisted over many months. We placed Delta-ORF7a^Δ17del^ within the context of all other ORF7a mutations that have occurred globally in Delta SARS-CoV-2 genomes, tested whether Delta-ORF7a^Δ17del^ produced a functional/detectable protein, determined where the ORF7a protein localises within infected cells, and examined *in vitro* growth of ORF7a^Δ17del^.

## Results

### Frequency of Delta-ORF7a^Δ17del^

Over 4 million (4,018,216) Delta SARS-CoV-2 consensus genomes downloaded from GISAID met inclusion criteria for characterisation of deletions in the ORF7a gene (see Materials and Methods). A total of 4,195 unique deletion patterns were detected, with 134,751/4,018,216 (3.35%) of genomes in the data set having some form of deletion in ORF7a (Table S1). Approximately 71% of ORF7a deletions were predicted to result in a frameshift, leading to a premature stop codon and truncated amino acid sequence. Most of the deletions were encountered infrequently (median: 2 genomes; IQR: 1-5 genomes), with only 19 different deletions occurring in greater than 1000 genome sequences (Table 1; Table S1). Focusing on genomes that were inferred to have a deletion in ORF7a, there was generally only one deletion detected in a given genome sequence (median: 1 deletion; IQR:1-1 deletions), but there was a maximum of six deletions found in three genomes. Generally, deletions were detected over short time periods (median: 1 day; IQR= 1-71 days). The longest-persisting in-frame deletion was a 12-nt deletion of bases 27617-27628 found in 99 genomes, which was first sampled on 2020-08-11 and last sampled on 2022-01-12, a time difference of 519 days (Table S1). Although frameshift-inducing deletions were also generally transient, as expected, some were sampled over a relatively long time period (Table 1; Table S1). For example, a 64-nucleotide frameshift-inducing deletion spanning positions 27556-27619 was found in 8945 genome sequences sampled between 2021-03-20 and 2022-02-22, a time difference of 339 days. Not surprisingly, most deletions were detected in only a few countries (median: 1 country; IQR 1-2 countries) with some evidence of geographical spread when limiting the analysis to deletions occurring in >1000 genomes (median 31; IQR 19-41 countries).

**Table 1.** The top 20 deletions in ORF7a of Delta genomes in GISAID clade GK (as of 2022-05-31), as ranked by the number of genomes in which they are found. Deletions are summarised in shorthand format in the form of (start coordinate, end coordinate, overall length in nucleotides). The deletion that characterised the Australian Delta outbreak (Delta-ORF7a^Δ17del^) is emphasised in bold. Accession numbers for all samples analysed for this table are in Table S1 and https://doi.org/10.55876/gis8.220809bs.

| Deletion | First Date Sampled | Last Date Sampled | Date Span (days) | Genomes | Countries | Frameshift-inducing |
| --- | --- | --- | --- | --- | --- | --- |
| **(27607, 27623, 17)** | **2021-06-02** | **2022-02-13** | **256** | **28014** | **27** | **TRUE** |
| (27556, 27619, 64) | 2021-03-20 | 2022-02-22 | 339 | 8945 | 17 | TRUE |
| (27579, 27581, 3) | 2021-01-22 | 2022-02-04 | 378 | 5189 | 68 | FALSE |
| (27551, 27577, 27) | 2021-02-23 | 2022-02-15 | 357 | 3109 | 30 | FALSE |
| (27692, 27697, 6) | 2021-03-09 | 2022-02-20 | 348 | 3108 | 54 | FALSE |
| (27538, 27628, 91) | 2021-06-04 | 2022-02-10 | 251 | 2887 | 44 | TRUE |
| (27588, 27606, 19) | 2021-06-22 | 2022-01-17 | 209 | 2655 | 8 | TRUE |
| (27555, 27578, 24) | 2021-05-19 | 2022-02-03 | 260 | 1916 | 51 | FALSE |
| (27720, 27721, 2) | 2021-05-12 | 2022-02-22 | 286 | 1859 | 46 | TRUE |
| (27553, 27621, 69) | 2021-04-09 | 2022-02-04 | 301 | 1636 | 33 | FALSE |
| (27554, 27621, 68) | 2021-04-12 | 2022-01-04 | 267 | 1585 | 21 | TRUE |
| (27694, 27700, 7) | 2021-02-09 | 2022-01-12 | 337 | 1538 | 38 | TRUE |
| (27548, 27623, 76) | 2021-06-21 | 2022-01-10 | 203 | 1418 | 13 | TRUE |
| (27548, 27554, 7) | 2021-01-11 | 2022-01-17 | 371 | 1290 | 31 | TRUE |
| (27578, 27623, 46) | 2021-04-04 | 2022-01-31 | 302 | 1135 | 34 | TRUE |
| (27555, 27624, 70) | 2021-04-05 | 2022-02-02 | 303 | 1115 | 28 | TRUE |
| (27564, 27586, 23) | 2021-06-09 | 2022-01-12 | 217 | 1044 | 17 | TRUE |
| (27553, 27553, 1) | 2021-04-22 | 2022-01-18 | 271 | 1007 | 35 | TRUE |
| (27555, 27633, 79) | 2021-07-26 | 2021-12-31 | 158 | 1003 | 7 | TRUE |
| (27695, 27700, 6) | 2021-04-16 | 2022-01-25 | 284 | 992 | 35 | FALSE |

By far the most common ORF7a deletion in the data set corresponds to genomic positions 27607–27623, as found in Delta-ORF7a^Δ17del^ genomes. Of the 134,751 Delta genomes in the filtered data set that have at least one deletion in ORF7a, 28014 (20.8%) and 166 (0.12%) possess ORF7a^Δ17del^ as either the sole deletion in ORF7a (Table 2) or as one of several deletions in the gene, respectively. The majority of Delta genomes with ORF7a^Δ17del^ in some form (27912/28180; 99.05%) originate from Australia (Table 2). Placing these genomes into context of circulating Delta viruses in Australia from 2021-06-14 (the official start date of the Delta outbreak) and last detected virus on the 2022-02-15, 87.1% (27912/32048) of Delta genomes originating from Australia possessed the ORF7a^Δ17del^ deletion (Figure 2; Table 2).

**Table 2.** All countries that generated genomes possessing the deletion that characterised the Australian Delta outbreak (ORF7a^Δ17del^) during the outbreak time period, as determined by analysis of all Delta genomes in GISAID clade GK (as of 2022-05-31). The genomes either possessed ORF7a^Δ17del^ as the sole deletion in ORF7a, or it was one of two or more deletions in the gene. The table is ranked by the number of genomes with ORF7a^Δ17del^. The percentage of Delta genomes submitted to GISAID during the time period that had ORF7a^Δ17del^ is given to two decimal places. Note: a date span of 0 days suggests that genome(s) were collected on a single date. Accession numbers for all samples analysed for this panel are in Table S1 and https://doi.org/10.55876/gis8.220809bs.

| Country | Genomes with ORF7a<sup>Δ17del</sup> Deletion | Total Genomes in Period | Percentage of Genomes with Deletion | First Date Sampled | Last Date Sampled | Date Span (days) |
| --- | --- | --- | --- | --- | --- | --- |
| Australia | 27912 | 32048 | 87.09 | 2021-06-13 | 2022-02-13 | 245 |
| USA | 102 | 1402035 | 0.01 | 2021-07-14 | 2021-12-20 | 159 |
| England | 76 | 865173 | 0.01 | 2021-06-02 | 2021-12-23 | 204 |
| Brazil | 24 | 40686 | 0.06 | 2021-09-20 | 2022-01-17 | 119 |
| France | 17 | 114469 | 0.01 | 2021-10-20 | 2021-12-27 | 68 |
| India | 7 | 73032 | 0.01 | 2021-09-18 | 2022-01-03 | 107 |
| Slovenia | 6 | 27823 | 0.02 | 2021-10-19 | 2021-12-10 | 52 |
| Thailand | 6 | 7775 | 0.08 | 2021-11-11 | 2021-12-19 | 38 |
| Netherlands | 4 | 44346 | 0.01 | 2021-11-16 | 2022-01-04 | 49 |
| Scotland | 4 | 103198 | 0.00 | 2021-10-28 | 2021-12-17 | 50 |
| Colombia | 2 | 4525 | 0.04 | 2021-11-22 | 2021-12-19 | 27 |
| Japan | 2 | 95139 | 0.00 | 2021-08-30 | 2021-09-28 | 29 |
| Mexico | 2 | 23422 | 0.01 | 2021-12-12 | 2021-12-12 | 0 |
| Romania | 2 | 5884 | 0.03 | 2021-09-30 | 2021-09-30 | 0 |
| Russia | 2 | 7647 | 0.03 | 2021-06-29 | 2021-11-02 | 126 |
| Bahrain | 1 | 1893 | 0.05 | 2021-11-11 | 2021-11-11 | 0 |
| Canada | 1 | 97883 | 0.00 | 2021-10-20 | 2021-10-20 | 0 |
| Chile | 1 | 8486 | 0.01 | 2021-11-28 | 2021-11-28 | 0 |
| Croatia | 1 | 14217 | 0.01 | 2021-09-04 | 2021-09-04 | 0 |
| Israel | 1 | 18019 | 0.01 | 2021-11-06 | 2021-11-06 | 0 |
| Latvia | 1 | 5220 | 0.02 | 2021-10-13 | 2021-10-13 | 0 |
| Lithuania | 1 | 15449 | 0.01 | 2021-10-30 | 2021-10-30 | 0 |
| Luxembourg | 1 | 9493 | 0.01 | 2021-11-29 | 2021-11-29 | 0 |
| Malta | 1 | 19 | 5.26 | 2022-01-08 | 2022-01-08 | 0 |
| Peru | 1 | 6575 | 0.02 | 2021-10-28 | 2021-10-28 | 0 |
| Vietnam | 1 | 2423 | 0.04 | 2021-11-16 | 2021-11-16 | 0 |
| Wales | 1 | 98861 | 0.00 | 2021-08-24 | 2021-08-24 | 0 |

**Figure 2.**
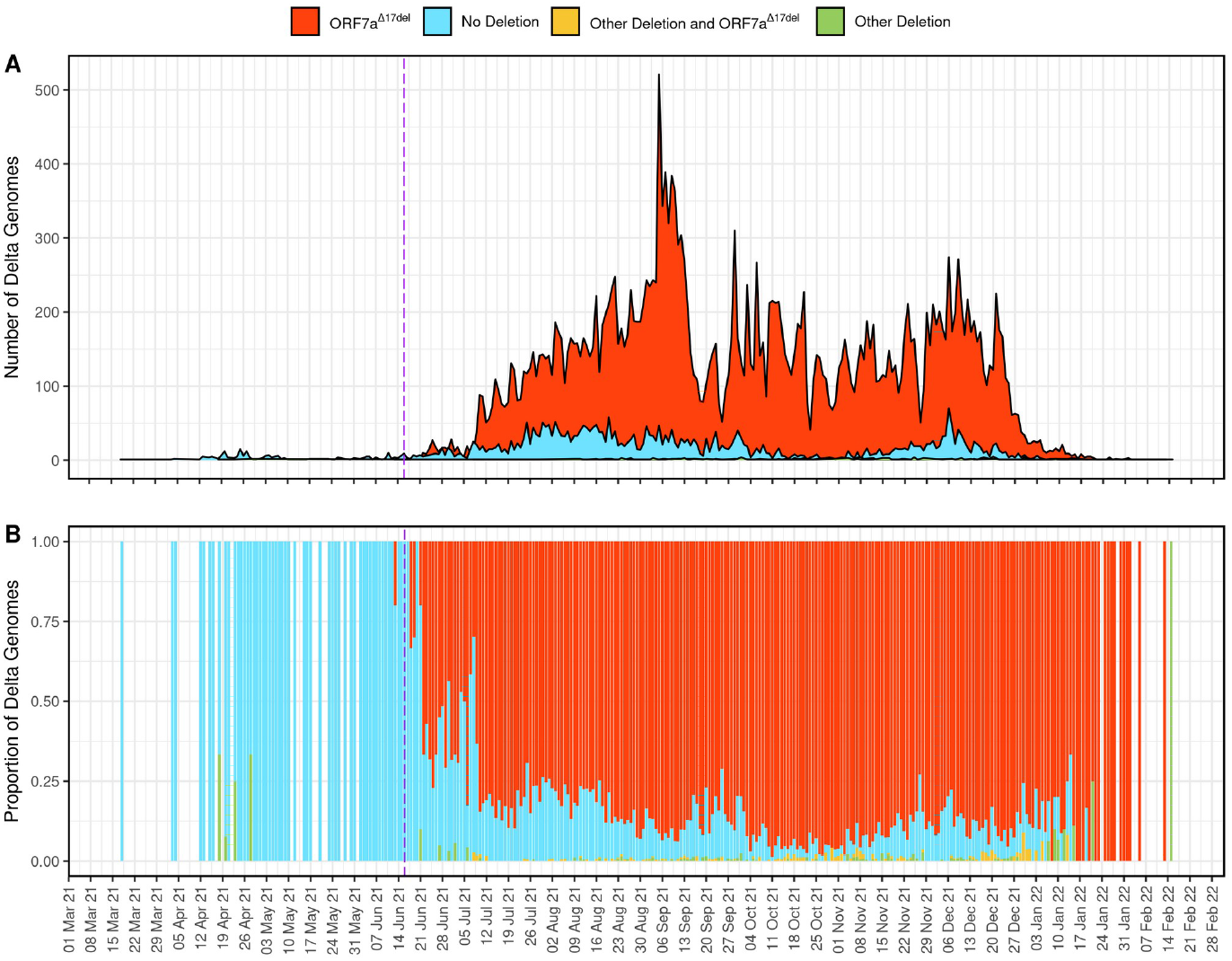
Time series plots of 32,048 genome sequences from the Australian Delta outbreak showing the number (**A**) and proportion (**B**) of genomes per day that possessed the characteristic deletion of 17 nucleotides within ORF7a spanning genomic positions 27607–27623 (ORF7a^Δ17del^), any other deletion in ORF7a, any other deletion in ORF7a plus ORF7a^Δ17del^, or no deletion in ORF7a. The purple dashed line indicates the beginning of the Australian Delta outbreak. Accession numbers for all samples analysed for this panel are in Table S1 and https://doi.org/10.55876/gis8.220809bs.

### Structure of ORF7a in Delta-ORF7a^Δ17del^

The Delta-ORF7a^Δ17del^ genome encodes for the first 70 amino acids of the ORF7a protein as normal. These amino acids include the signal peptide, which is retained in all SARS-CoV-2 ORFs and assists with protein secretion, followed by 6 beta sheets (Figure 3A). The 17-nt deletion characteristic of Delta-ORF7a^Δ17del^ genomes occurs after the 6th beta sheet, leading to 7 novel amino acids followed by a stop codon (Figure 1B; Figure 3B), and causing the loss of the 7th beta sheet, the C-terminal transmembrane domain and the cytoplasmic di-lysine motif (KRKTE) that determines ER localisation. In addition, it also leads to the loss of the ORF7a:K119 amino acid, which is polyubiquitinated and is thought to lead to IFN-I inhibition via blocking of STAT2 (8, 9). The loss of the 50 C-terminal residues is predicted to have a major impact on function. Firstly, the loss of the ORF7a:K119 amino acid is expected to cause the loss of IFN-I suppression in Delta-ORF7a^Δ17del^, resulting in reduced replication titres. Secondly, as ORF7a is normally retained on the cell surface and intracellularly on the Golgi and ER membrane, this deletion along with the retention of the N-terminal signal peptide might result in increased secretion of this protein. Given that the ORF7a ectodomain has been shown to interact with CD14+ monocytes with high efficiency, an increased secretion of this protein systemically could result in an altered pathology of this variant (6). Although protein docking analysis indicated that the ORF7a protein of Delta-ORF7a^Δ17del^ could bind CD14, the loss of the 7^th^ beta sheet and the K119 significantly altered the proposed interaction sites (Table S2).

**Figure 3.**
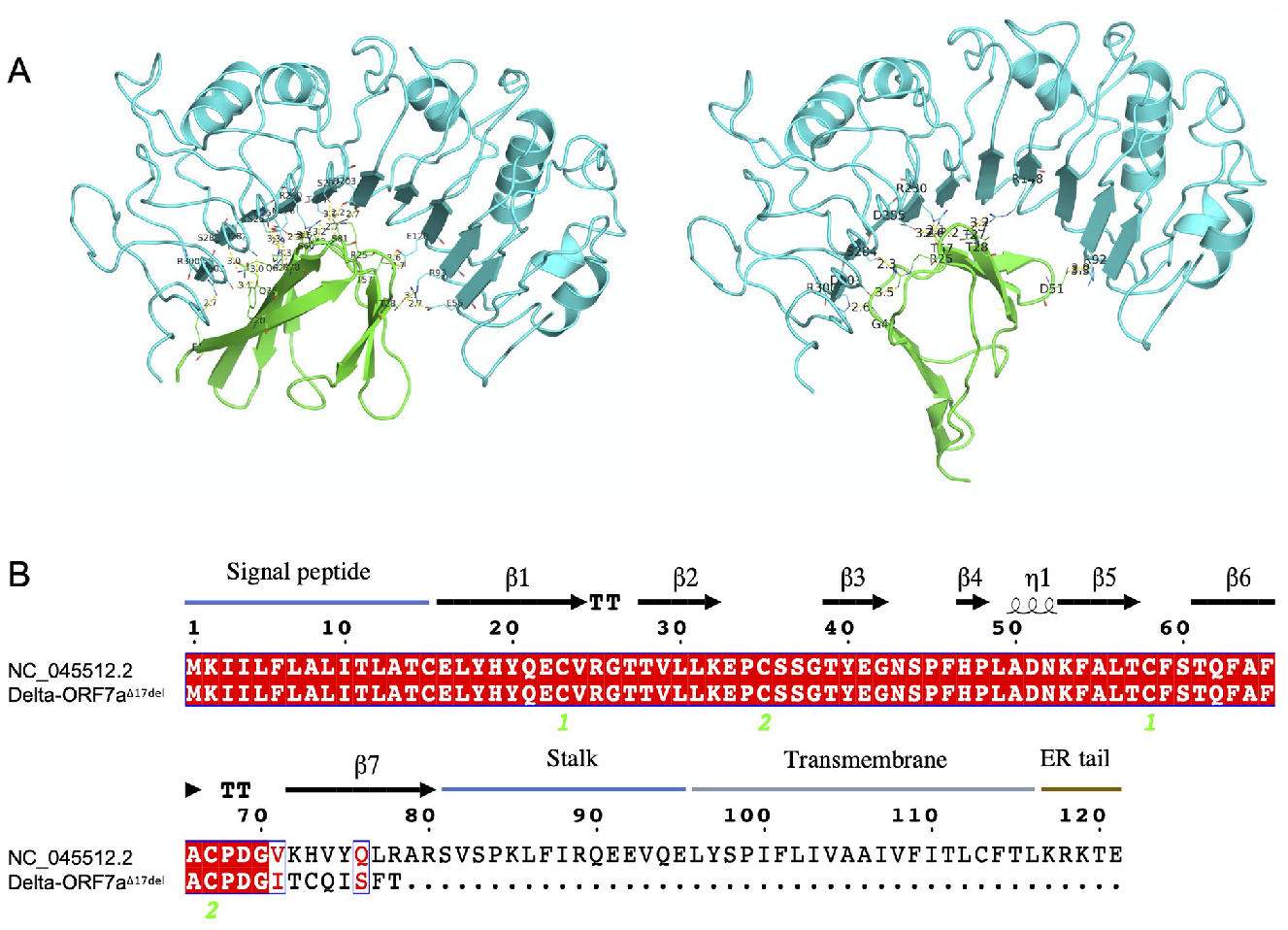
The estimated structure of the ORF7a protein in SARS-CoV-2, and the consequences of a deletion of 17 nucleotides within ORF7a spanning genomic positions 27607–27623 (ORF7a ^Δ17del^) on this structure. **A**. The ORF7a protein from the Wuhan-Hu-1 reference genome (PDB ID: 7CI3) docking with human CD14 (PDB ID: 4GLP), as modelled using HDOCK (32). **B**. Inferred peptide sequence of the ORF7a protein in SARS-CoV-2, demonstrating the peptide and structural changes within the Delta-ORF7a ^Δ17del^ variant. The ORF7a^Δ17del^ deletion occurs after the 6th beta sheet in Delta-ORF7a^Δ17del^, leading to 7 novel amino acids followed by a stop codon, and causing the loss of the 7th beta sheet, the C-terminal transmembrane domain and the cytoplasmic di-lysine motif (KRKTE) that determines ER localisation. The alignment was created using Esprit 3.0 (30).

### *In vitro* infection of Calu-3 cells with different SARS-CoV-2 variants

Cell lines were infected with an early A.2.2 variant of SARS-CoV-2, Delta-ORF7a^intact^, or Delta-ORF7a^Δ17del^. Western blots of the cell supernatants of uninfected cells and cells infected with the

A.2.2 variant of SARS-CoV-2, Delta-ORF7a^intact^, or Delta-ORF7a^Δ17del^ showed that nucleocapsid and spike protein was detected in the supernatant from all three variants, but not from uninfected cells as expected. Conversely, while ORF7a protein was detected in the supernatants of the A.2.2 and Delta-ORF7a^intact^, it was not detected in the supernatant for the Delta-ORF7a^Δ17del^ variant, indicating that it is not secreted in the latter variant (Figure S1).

Immunofluorescence staining used to compare the *in vitro* viral growth showed that 78.9% ± 3.6% cells infected with Delta-ORF7a^Δ17del^, 61.1% ± 4.2% of cells infected with Delta-ORF7a^intact^, 66.5% ± 6.9% of cells infected with the A.2.2 variant of SARS-CoV-2 and none of the uninfected cells were positive for the Nucleocapsid protein (Figure 4). This higher proportion Nucleocapsid protein expression in the Delta-ORF7a^Δ17del^ infected cells was statistically significant when compared to those infected with Delta-ORF7a^intact^ (p=0.008; unpaired test) but was not significant when compared to cells infected with the A.2.2 variant (p=0.183) (Figure 4).

**Figure 4.**
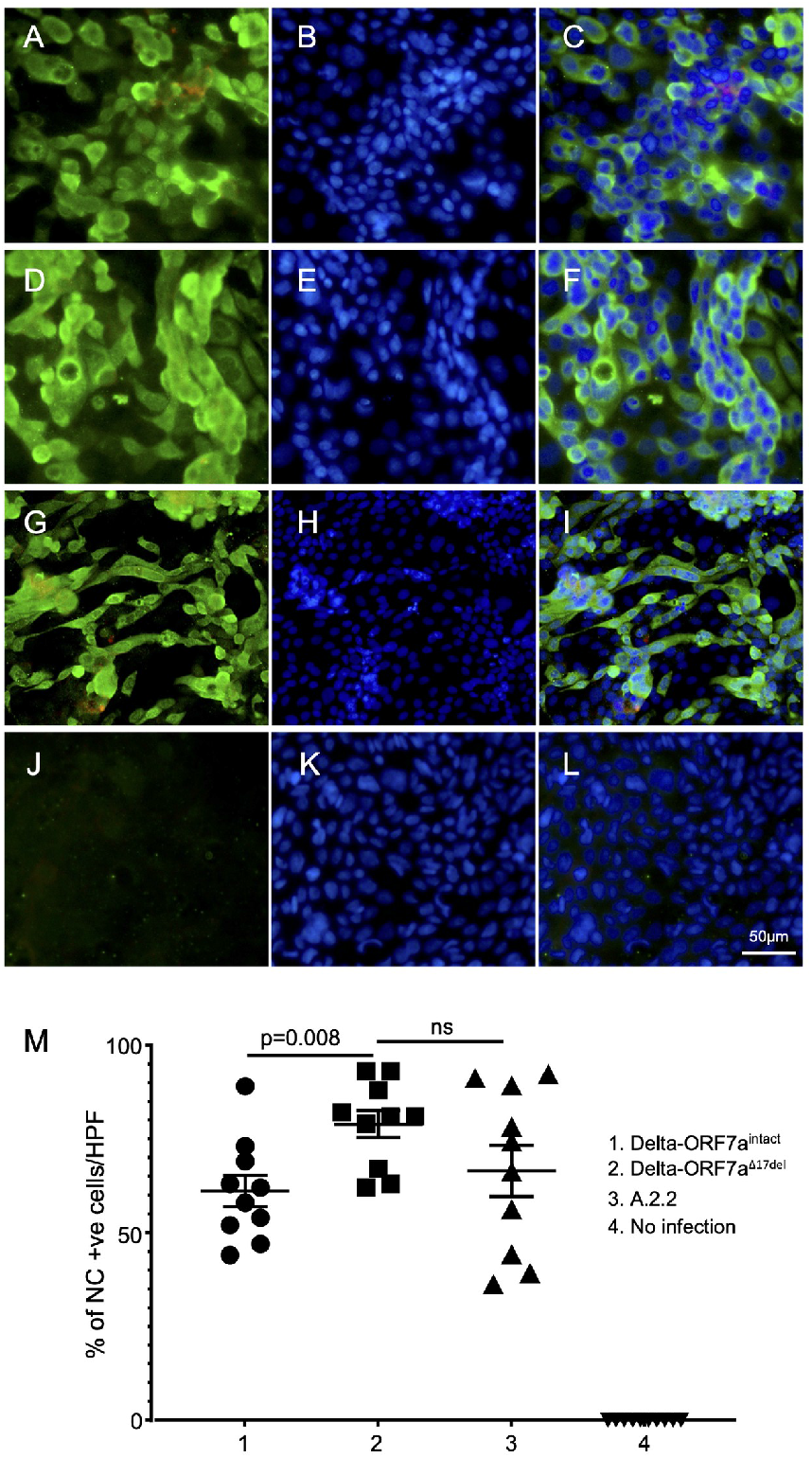
Nucleocapsid immunofluorescence staining of Calu-3 cells infected with SARS-CoV-2 variants. The panels in each row (**A-C, D-F, G-I, J-L**) show images of cells infected with Delta ORF7a^intact^, Delta ORF7a^Δ17del^, the A.2.2 variant, and no infection, respectively. The panels in each column show cells with green Alexa-488 fluorescence staining to detect SARS-CoV-2 Nucleocapsid, blue DAPI nuclear staining, and merged Alexa-488 and DAPI staining, respectively. There is abundant cytoplasmic expression of Nucleocapsid protein in all infected cells, but none in uninfected cells. **M**. Summary plot showing more proportions of Nucleocapsid positive cells in those infected with Delta-ORF7a^Δ17del^ compared to Delta-ORF7a^intact^, A.2.2 variant infected cells, and uninfected cells (n=10 high power fields per group; mean ± SEM; p values calculated by two tailed unpaired t tests).

The mean fluorescent intensity (MFI) staining of the ORF7a protein in Delta-ORF7a^Δ17del^ infected cells was 70.7 ± 0.98, which was significantly less than the MFI staining of those cells infected with the A.2.2 variant (74.4 ± 0.95; p=0.004, unpaired t test) (Figure 5). Moreover, the expression pattern of the protein in the A.2.2-variant-infected cells was discrete throughout the cytoplasm, with some nuclear staining, while in Delta-ORF7a^Δ17del^-infected cells the expression pattern was mostly diffused throughout the cytoplasm. Delta-ORF7a^intact^ cells were not available for ORF7a staining. No ORF7a protein was detected in the uninfected cells (Figure 5).

**Figure 5.**
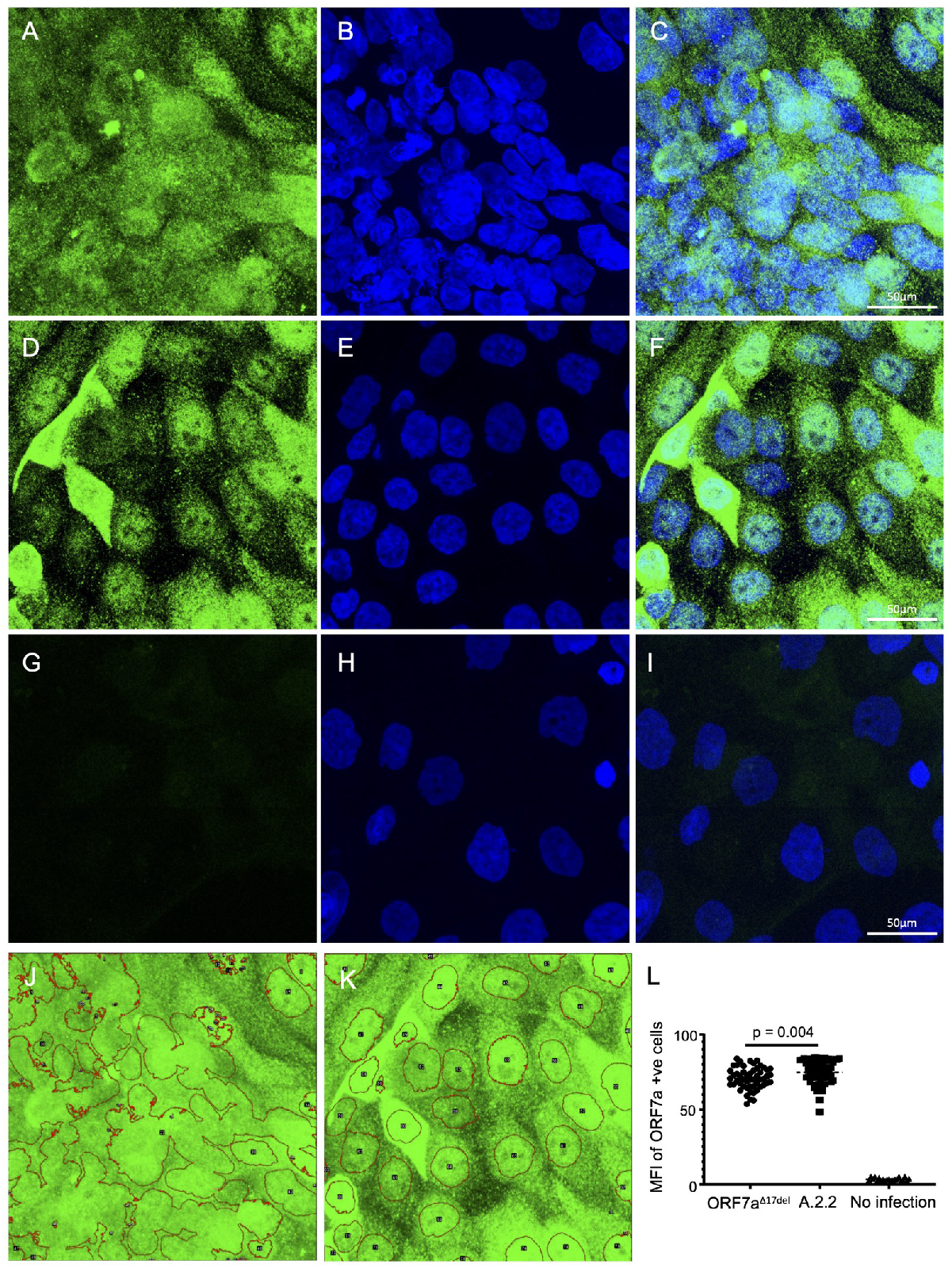
ORF7a immunofluorescence staining of Calu-3 cells infected with SARS-CoV-2 variants. **A, B, C**. Cells infected with Delta-ORF7a^Δ17del^ showing diffused cytoplasmic and nuclear staining (green, A) with DAPI nuclear staining (blue, B) and merged image (C). **D, E, F**. Cells infected with the A.2.2. variant showing abundant and discrete cytoplasmic and some nuclear staining (green, D) with DAPI nuclear staining (blue, E) and merged image (F). **G, H, I**. No ORF7a staining in uninfected cells. **J, K**. Representative NIH Image J mean fluorescence analysis of individual cells infected with Delta ORF7a^Δ17del^ (J) and the A.2.2. variant (K). **L**. Summary plot of the mean fluorescent intensity of ORF7a positive cells in those infected with Delta-ORF7a^Δ17del^ (“ORF7a^Δ17del^”) compared to the A.2.2 variant infected cells and uninfected cells (21-78 cells were analysed per group; mean ± SEM; p value calculated by two tailed unpaired t tests).

## Discussion

Deletions in the ORF7a region of SARS-CoV-2 have been documented even since early in the COVID-19 pandemic (2, 3, 10–13). Our analysis of >4 million genomes from clade GK of the SARS-CoV-2 phylogeny demonstrates that ORF7a remains a region where deletions continue to occur, with 3.35% of genomes possessing a deletion in ORF7a (Table S1). Previous studies from the COVID-19 pandemic documented ORF7a variants including those that induce frameshifts (2), with evidence of frequent independent emergence. Sustained transmission of these SARS-CoV-2 variants was encountered infrequently, likely secondary to the reduced viral fitness of these variants (2). Our results support these findings: 4194 unique ORF7a deletions were detected in our data set of clade GK genomes, but many of these were only sampled once on a single date of collection. Clearly, ORF7a remains a hotspot for deletions in the SARS-CoV-2 genome, and the reason for this phenomenon should continue to be investigated.

Unlike previous studies conducted earlier in the COVID-19 pandemic, we show that some ORF7a variants can persist over long time periods, in some cases longer than one year (Table 1; Table S1). However, these genomes with ORF7a deletions seem to be predominantly sampled at low frequencies relative to the dominant circulating strain. The Australian Delta outbreak is therefore clearly unique for several reasons. First, the dominant circulating subtype was an ORF7a variant (Delta-ORF7a^Δ17del^; deletion of genomic positions 27607–27623; Figure 2), accounting for ∼87% of Delta genomes in Australia at the time. Second, there was ongoing transmission of this distinctive deletion, with 27,912 genomes sampled over ∼7 months (245 days). The true dominance of Delta-ORF7a^Δ17del^ genomes in Australia could even be underestimated here since the data set includes returned travellers to Australia (more likely to lack the mutation characteristic of Australian cases), and genomes with potentially incomplete ORF7a sequences (i.e., those potentially lacking coverage at some or all of positions 27607–27623). Additionally, variant calling software can have difficulties with correctly calling insertion/deletions (indels) (14), and by default GISAID does not release submissions that contain a frameshift mutation (unless specifically requested). Finally, the ongoing transmission of Delta-ORF7a^Δ17del^ occurred despite cocirculation of presumably a more “fit” virus (Delta-ORF7a^intact^). Truncation of the ORF7a C-terminus, leading to loss of the transmembrane domain, has been associated with decreased viral fitness in some (but not all) studies by negating any anti-immune properties of the protein (15).

There are two main possibilities that might explain the persistence and success of Delta-ORF7a^Δ17del^ in Australia. Firstly, the 17-nt deletion could have become (near-)fixed in the outbreak population by chance alone during an initial transmission bottleneck and persisted despite a fitness disadvantage due to a lack of competition from Delta-ORF7a^intact^ viruses and/or other circulating subtypes at that time. Alternatively, the ORF7a^Δ17del^ deletion was not deleterious to the virus, thereby allowing Delta-ORF7a^Δ17del^ variants to persist and proliferate.

Given the unique properties and prolonged persistence of Delta-ORF7a^Δ17del^, we sought to determine whether Delta-ORF7a^Δ17del^ viruses might have a potential fitness advantage compared to other contemporaneous Delta viruses lacking the ORF7a^Δ17del^ deletion (Delta-ORF7a^intact^) and/or an ancestral lineage of SARS-CoV-2 (A.2.2). We used infection of cells *in vitro* as a proxy for a potential fitness advantage. We posited that the frameshift mutation might cause a difference in cell localisation of the ORF7a protein because of a loss of the transmembrane domain, which has been shown to lead to localisation of the ORF7a at the golgi, ER and cell surface (1).

Despite infecting with the same multiplicity of infection (MOI) of A.2.2 and Delta-ORF7a^intact^ variants, the staining for nucleocapsid in infected cells suggested that the Delta-ORF7a^Δ17del^ variant was replicating to a higher titre than the Delta-ORF7a^intact^ variant, but similar to the A.2.2 variant. However, staining for ORF7a showed that this protein was generated at a lower abundance in the Delta-ORF7a^Δ17del^ infected cells, and it appeared to be mostly diffused in the cell cytoplasm, as opposed to A.2.2, where ORF7a was also stained within cell nuclei. One would expect this reduced amount of ORF7a protein to, at best, not lead to increased viral fitness, and, at worst, be detrimental to fitness. Unfortunately Delta-ORF7a^intact^ was not available for ORF7a staining and cannot be compared here.

Since the truncated ORF7a protein in Delta-ORF7a^Δ17del^ lacks a transmembrane domain, increased secretion of the truncated ORF7a protein in the supernatant relative to the other variants could be expected. However, ORF7a was not detected in the supernatant of the Delta-ORF7a^Δ17del^ variant by western blot, although this result could be a sensitivity issue given the lower level of intracellular ORF7a observed in culture. Overall, we do not see attenuated growth of Delta-ORF7a^Δ17del^ in comparison to the other tested variants, which was unexpected since expression of functional ORF7a has been reported to inhibit BST2, leading to enhanced virion production (Wang et al. 2014). A possible explanation is that an ability to bind to BST2 was retained in Delta-ORF7a^Δ17del^ since all but one of the proposed binding sites to BST2 (L17, H19, Q20, D69 and R80) are retained in the truncated protein (4).

In summary, the *in vitro* results do not suggest any overt fitness disadvantage or advantage of Delta-ORF7a^Δ17del^ compared to Delta-ORF7a^intact^ in cell culture. No strong fitness advantage from the ORF7a^Δ17del^ mutation is to be expected given the generally deleterious effect of frameshift mutations leading to partial or full loss of gene function (16). Despite this, Delta-ORF7a^Δ17del^ were overwhelmingly the infecting virus in COVID-19 within Australia during the outbreak period. The most likely reason for this is an initial founder effect leading to a lack of competition. At the beginning of the Delta outbreak, there was no community transmission of SARS-CoV-2 in NSW, and very little transmission in other regions of Australia. Previous instances of community transmission were restricted to relatively small, isolated clusters, and widespread vaccination was yet to be achieved. Consequently, much of the Australian population was immunologically naive to SARS-CoV-2. Additionally, multiple competing variants were not introduced into the community concurrently, partly resulting from strong border controls. These conditions were ideal for the spread of a virus bearing a mutation of some (mild) fitness benefit, that otherwise might be outcompeted in the presence of other lineages (17). Indeed, different geographic regions have had isolated outbreaks of SARS-CoV-2 with distinct mutational profiles as a result of founder effects even since early in the pandemic (18). These characteristic mutations are useful for genomic epidemiology by allowing sequences to be assigned to a given cluster or geographic region (19). It is also important to note that, even when not causing an increased infectivity or severity, mutations can still impact genomic epidemiology by affecting the performance of amplicon schemes of qPCR diagnostics (e.g., 20).

## Conclusion

The rapid proliferation of a particular mutation in the genome of a virus during an outbreak(s) can often trigger concern. Often this concern turns out to be warranted, such as in cases where mutations clearly lead to increased infectivity and/or immune evasion (21–23). However, the phenomena of genetic bottlenecks and founder effects clearly demonstrate that a sudden dominance of a given mutation does not necessarily imply positive selection leading to a more dangerous virus (24). Given the lack of a strong difference in *in vitro* growth between Delta-ORF7a^Δ17del^ and a contemporaneous ‘wild type’ Delta virus, the most likely conclusion is that the Australian Delta outbreak was not made any worse by the presence of the characteristic ORF7a^Δ17del^ mutation. However, we demonstrate that ORF7a deletions have continued to occur relatively frequently in viruses from many countries throughout the pandemic, echoing observations from early studies. This trend implies that there might still exist some benefit of deletions in ORF7a to SARS-CoV-2, such as by somehow compensating for more deleterious mutations elsewhere in the genome (25). Further research into deletions in ORF7a might prove useful to understand the ongoing evolution of SARS-CoV-2, in addition to providing useful markers for genomic epidemiology.

## Materials and Methods

### Analysis of deletions

GISAID employs a nomenclature system whereby clades in the global outbreak tree are named based on the presence of key defining mutations. Clade GK predominantly comprises Delta genomes; therefore, we downloaded all glade GK genomes available as a direct download from GISAID (4,223,396 genomes as of 2022-05-31). The full list is available via http://gisaid.org/EPI_SET_220809bs or https://doi.org/10.55876/gis8.220809bs, and the filtered data set is within Table S1.

We searched each genome in the dataset for the presence of any deletions within the ORF7a gene using ‘deletion_detector’, a custom program written in python3 for the purposes of this study (26). Briefly, this program aligns each input sequence against the Wuhan Hu-1 reference genome for SARS-CoV-2 using minimap2 v2.24 (27), converts the SAM-format alignment into fasta format using gofasta v1.1.0 (28), then uses custom python code to find the coordinates and lengths of all gaps in a region of the genome of interest. The output file (TSV format) contains the genomic coordinates of deletions within query sequences relative to the reference genome as well as other quality control metrics, depending on the input parameters to the analysis. The date and country of collection for query sequences are automatically parsed from GISAID headers. All analyses are conducted efficiently using parallel processing of input query files. The ‘deletion_detector’ program is freely available as open-source software from Zenodo (26) or GitHub (https://github.com/charlesfoster/deletion_detector).

Clade GK also comprises some other lineages that have independently acquired the defining mutations of the clade. To restrict our survey of ORF7a deletions to only global Delta genomes, we assigned Pango lineages to all downloaded genomes using pangolin v4.06 with pangoLEARN v1.9 (29). We then imported the results from ‘deletion_detector’ into R and filtered out all samples that were not assigned to the Delta lineage, were known duplicates, were missing coverage for the entirety of ORF7a, and/or did not have a complete date of collection (YYYY-MM-DD). After this filter, 4,018,216 genomes remained. Summary statistics were then estimated and plotted in R.

### Structural Modeling

Protein alignment was performed with Esprit3.0 (30). For the tertiary structural modelling of the ORF7a protein of SARS-CoV-2, the SWISS-MODEL software (31) was used with the resolved structure of the ORF7a protein from the SARS-CoV-2 Wuhan-Hu-1 variant (PDB ID: 7CI3) input as template. The model had a Molprobity score of 1.17 and Ramachandran favored 96.55%. To investigate the potential binding interfaces of SARS-COV-2 Delta ORF7a with CD14+ monocytes, we performed molecular docking experiments using HDOCK (32). The structure of human CD14 antigen was obtained from the RCSB website (PDB ID: 4GLP). Top 10 interactions provided by the software were observed manually and the model with the highest score and best interaction was selected (Figure 3). The model was further visualized by using Pymol and interaction between the amino-acid residues were calculated.

### Infection of Calu-3 cell line

Calu-3 cells were maintained in minimum essential media (MEM; Sigma, M4655) containing 10% fetal bovine serum (FBS; Sigma, F423-500) and 1x penicillin-streptomycin (Gibco, 15140122). Cells were seeded in a 24-well tissue culture plate containing plasma-cleaned-PLL-coated coverslips (3×10^5^ cells/well) and incubated for 24 hours. Maintenance media was aspirated, and wells were infected in quadruplicate with 10^3^ TCID_50_ of Ancestral (A.2.2), Delta wild-type (B.1.617.2), and Delta-ORF7a^Δ17del^ variants of SARS-CoV-2 (Table 3). Cultures were incubated for 48 hours (37°C, 5% CO_2_) before supernatants were harvested and inactivated with beta-propiolactone for western blot analysis, and cells were fixed with paraformaldehyde for immunofluorescent staining.

**Table 3.**
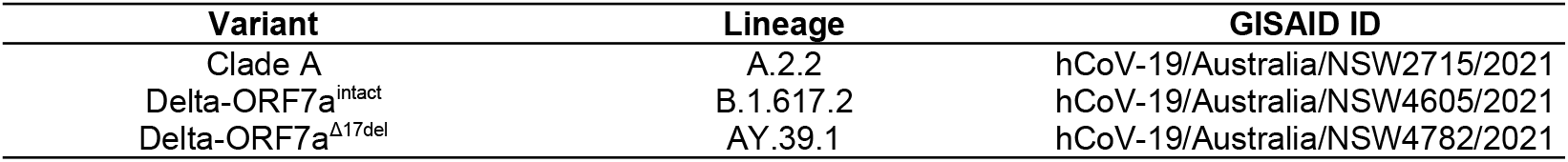
Variants used in Calu-3 infection

| Variant | Lineage | GISAID ID |
| --- | --- | --- |
| Clade A | A.2.2 | hCoV-19/Australia/NSW2715/2021 |
| Delta-ORF7a<sup>intact</sup> | B.1.617.2 | hCoV-19/Australia/NSW4605/2021 |
| Delta-ORF7a<sup>Δ17del</sup> | AY.39.1 | hCoV-19/Australia/NSW4782/2021 |

### Western blot

Cell supernatants were concentrated with the Amicon 8050 using a 10kDa membrane following manufactures protocol (UFSC05001, Millipore) and 10ul loaded onto 4-20% SDS-PAGE for western blot. The SDS-PAGE was resolved at 200 volts for ∼30 minutes, until the dye front reached the bottom of the gel. It was then transferred to Immobilon-P PVDF Membrane (IPVH0010, Merck) for 1hr at 400 mA. Western blots were blocked with 5% Skim Milk (PBS-T) for 1hr at room temperature then assessed with either 1:1000 anti-SARS-CoV-2 Nucleocapsid (NB100-56576, Novus Bio), 1:1000 anti-beta actin A5316, Sigma), 1:1000 anti-ORF7a mouse-MAb (ab273435, Abcam) or 1:1000 anti-SARS-CoV-2 Spike rabbit-MAb (40150-T62, Sino Biological) and incubated overnight at 4ºC. Following day the blot was washed three times in PBS-T, followed by a 2hr room temperature incubation of either goat anti-mouse HRP (172-1011, BioRad) or goat anti-rabbit HRP (170-6515, BioRad) at 1:5000 and 1:3000, respectively. After another three washes in PBS-T, the blots were developed with Clarity Max Western ECL Substrate (170-5062, BioRad) for 5 minutes then imaged using the G:box Chemi XX6 (Syngene).

### Immunofluorescent staining

Calu-3 cells infected with A.2.2, Delta and Delta-ORF7a^Δ17del^ or SARS-CoV-2 infected and non-infected Calu-3 controlscells on plasma-cleaned-PLL-coated coverslips were washed once with PBS and fixed with 4% paraformaldehyde for 10 minutes at room temperature (RT) then rinsed 3 times with PBS. The auto-fluorescence of paraformaldehyde was quenched with 50 mM ammonium chloride in PBS for 10 minutes at RT. After 3 washes with PBS, cells were permeabilized with 0.1% Triton X-100 in PBS for 5 minutes at RT, blocked with blocking buffer (0.02% Tween-20, 2% FBS in PBS) for 30 minutes at RT and then incubated with mouse anti-ORF7a IgG1 mAb (ab273435) (Abcam,), rabbit anti-nucleocapsid polyclonal IgG (NB 100-56576) (Novus Biologicals, CO, USA) or isotype matched mouse IgG1 mAb negative control or irrelevant rabbit IgG control (Sigma, Australia) (all used at 1 μg/ml diluted in the blocking buffer) overnight at 4°C. Next day, cells were washed 4 times with PBS and incubated with Alexa 488-conjugated goat anti-mouse or goat anti-rabbit Ab (1:400 dilution in PBS with 0.1% Triton X-100; Molecular Probes, VIC, Australia) for 1.5 hours at RT, washed 4 times with PBS and mounted with DAPI containing media (ProLong, Thermo Fisher) The percentages of nucleocapsid positive cells in 10 randomly selected fields at 250X magnification imaged by Olympus DP74 fluorescent microscope were counted using NIH Image J version 1.52. The ORF7a stained cells were imaged using Zeiss LSM 880 laser scanning confocal microscope at 650X magnification and the average fluorescent intensity (MFI) of all cells measured using NIH Image J version 1.52. MFI was measured in 21 uninfected cells, 53 Delta-ORF7a^Δ17del^ infected cells and 78 A.2.2 infected cells.

## Data Availability

All consensus genomes analysed in this study were downloaded from GISAID. The full list of accessions and associated metadata is available via http://gisaid.org/EPI_SET_220809bs or https://doi.org/10.55876/gis8.220809bs, and the list of accessions for genomes in our filtered data set upon which our conclusions are based is within Table S1. The custom program designed for the purposes of analysis in this study is freely available from https://github.com/charlesfoster/deletion_detector or https://doi.org/10.5281/zenodo.7049120. All other data are contained within the manuscript and its supplementary files.

https://dx.doi.org/10.6084/m9.figshare.21012535

## Acknowledgments

We gratefully acknowledge all data contributors, including the Authors and their Originating laboratories responsible for obtaining the specimens, and their Submitting laboratories for generating the genetic sequence and metadata and sharing via the GISAID Initiative, on which a component of this research is based. We thank the Royal College of Pathologists of Australasia Quality Assurance Programs (RCPAQAP), Medical Research Future Fund (MRFF) and the National Health and Medical Research Council (NHMRC) for financial support (COVID-19 Genomics Grant MRF9200006).

## Author contributions

The initial study was conceived and designed by C.S.P.F. and W.D.R., with contributions by R.A.B. and N.T. Infection of cell lines was managed by G.W., R.A.B., and D.A. Recombinant ORF7a protein production and Western blotting was carried out by F.S. and D.A. Immunofluorescence staining, imaging and analysis of the resulting data, was done by A.A. and N.T. The structure of ORF7a was modelled by R.A.B. and L.B.S. All other bioinformatic analysis was carried out by C.S.P.F. The manuscript was drafted by C.S.P.F., with all co-authors providing feedback in revisions and discussions, and approving the final version.

## Figures

**Figure S1.**
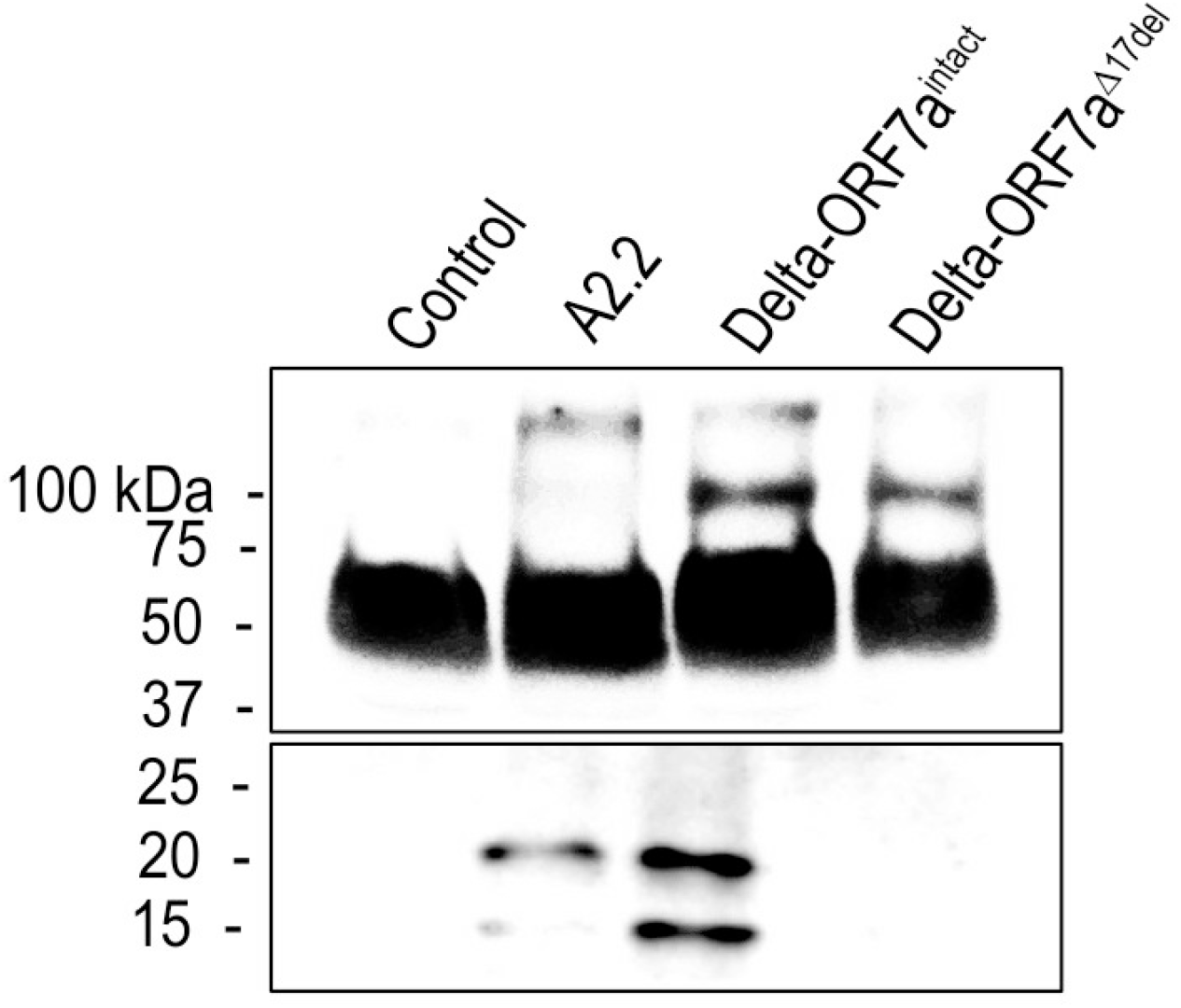
Western blot to detect ORF7a protein in the supernatant from cell lines infected with an early A.2.2 variant of SARS-CoV-2, Delta-ORF7a^intact^, or Delta-ORF7a^Δ17del^.

---

**Table S1.** All inferred deletions in the ORF7a region, and associated quality control metrics, of 4,018,216 Delta SARS-CoV-2 clade GK genomes downloaded from GISAID on 2022-05-31. Note: Table is too large for this document and can be accessed from:

- doi: 10.6084/m9.figshare.21012535 (reserved DOI that becomes public upon data publication)

**Table S2.**
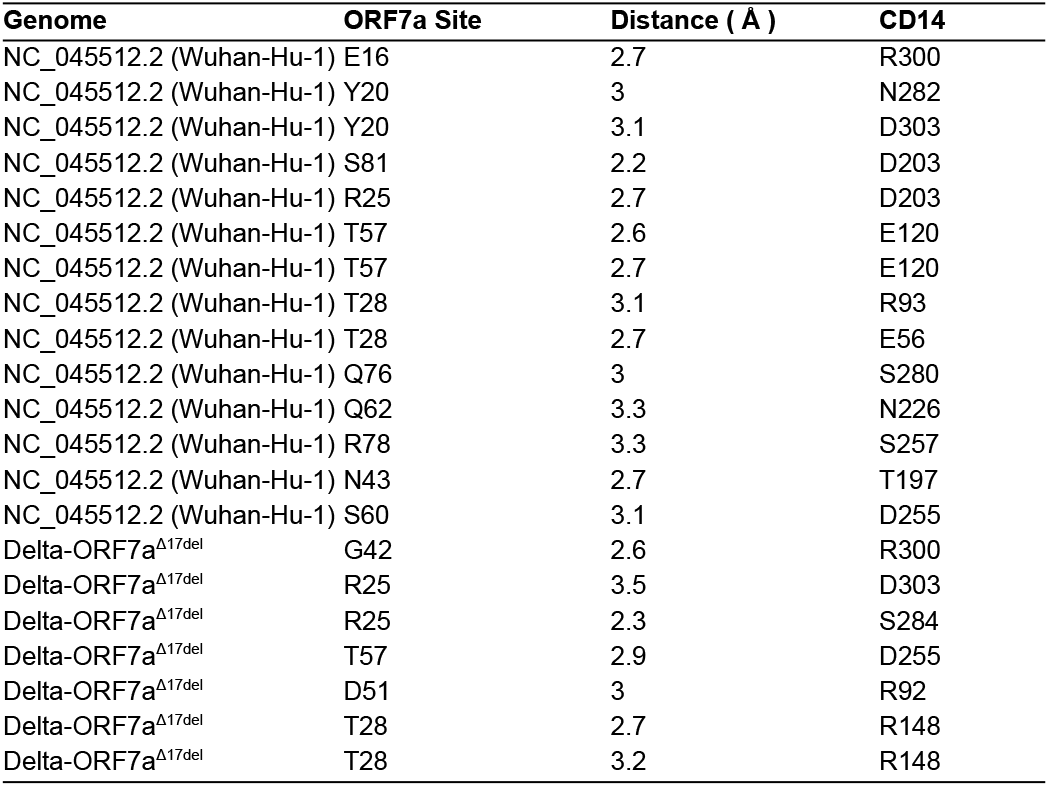
Inferred binding sites between SARS-CoV-2 ORF7a and human CD14. The inferences were made using HDOCK based on the ORF7a sequence from the Wuhan-Hu-1 reference genome and from the Delta-ORF7a^Δ17del^ variant.

