## Supplementary figures and images for "Persistence of a SARS-CoV-2 variant with a frameshifting deletion for the duration of a major outbreak"

### Figure S1

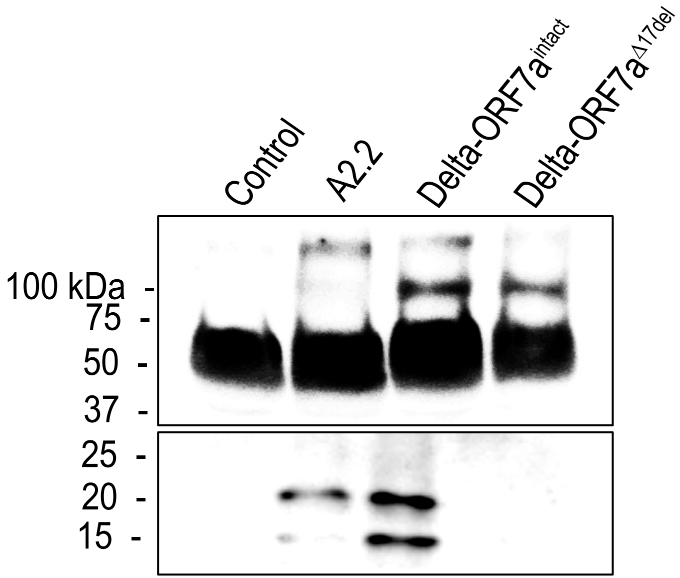
