## Supplementary material for "Persistence of a SARS-CoV-2 variant with a frameshifting deletion for the duration of a major outbreak": Table S2

**Table S2:** Inferred binding sites between SARS-CoV-2 ORF7a and human CD14. The inferences were made using HDOCK based on the ORF7a sequence from the Wuhan-Hu-1 reference genome and from the Delta-ORF7a^Δ17del^ variant.

| **Genome** | **ORF7a Site** | **Distance ( Å )** | **CD14** |
| --- | --- | --- | --- |
| NC_045512.2 (Wuhan-Hu-1) | E16 | 2.7 | R300 |
| NC_045512.2 (Wuhan-Hu-1) | Y20 | 3 | N282 |
| NC_045512.2 (Wuhan-Hu-1) | Y20 | 3.1 | D303 |
| NC_045512.2 (Wuhan-Hu-1) | S81 | 2.2 | D203 |
| NC_045512.2 (Wuhan-Hu-1) | R25 | 2.7 | D203 |
| NC_045512.2 (Wuhan-Hu-1) | T57 | 2.6 | E120 |
| NC_045512.2 (Wuhan-Hu-1) | T57 | 2.7 | E120 |
| NC_045512.2 (Wuhan-Hu-1) | T28 | 3.1 | R93 |
| NC_045512.2 (Wuhan-Hu-1) | T28 | 2.7 | E56 |
| NC_045512.2 (Wuhan-Hu-1) | Q76 | 3 | S280 |
| NC_045512.2 (Wuhan-Hu-1) | Q62 | 3.3 | N226 |
| NC_045512.2 (Wuhan-Hu-1) | R78 | 3.3 | S257 |
| NC_045512.2 (Wuhan-Hu-1) | N43 | 2.7 | T197 |
| NC_045512.2 (Wuhan-Hu-1) | S60 | 3.1 | D255 |
| Delta-ORF7a^Δ17del^ | G42 | 2.6 | R300 |
| Delta-ORF7a^Δ17del^ | R25 | 3.5 | D303 |
| Delta-ORF7a^Δ17del^ | R25 | 2.3 | S284 |
| Delta-ORF7a^Δ17del^ | T57 | 2.9 | D255 |
| Delta-ORF7a^Δ17del^ | D51 | 3 | R92 |
| Delta-ORF7a^Δ17del^ | T28 | 2.7 | R148 |
| Delta-ORF7a^Δ17del^ | T28 | 3.2 | R148 |
